# Two new compartmental epidemiological models and their equilibria

**DOI:** 10.1101/2021.09.03.21263050

**Authors:** Jonas Balisacan, Monique Chyba, Corey Shanbrom

**Affiliations:** UNIVERSITY OF HAWAI’I AT MĀNOA, 2565 MCCARTHY MALL, HONOLULU, HI 96822, USA; CALIFORNIA STATE UNIVERSITY, SACRAMENTO, 6000 J ST., SACRAMENTO, CA 95819, USA

**Keywords:** Compartmental model, reproductive number, endemic equilibrium, endemic threshold

## Abstract

Compartmental models have long served as important tools in mathematical epidemiology, with their usefulness highlighted by the recent COVID-19 pandemic. However, most of the classical models fail to account for certain features of this disease and others like it, such as the ability of exposed individuals to recover without becoming infectious, or the possibility that asymptomatic individuals can indeed transmit the disease but at a lesser rate than the symptomatic. Furthermore, the rise of new disease variants and the imperfection of vaccines suggest that concept of endemic equilibrium is perhaps more pertinent than that of herd immunity.

Here we propose a new compartmental epidemiological model and study its equilibria, characterizing the stability of both the endemic and disease-free equilibria in terms of the basic reproductive number. Moreover, we introduce a second compartmental model, generalizing our first, which accounts for vaccinated individuals, and begin an analysis of its equilibria.

## 1. Introduction

When modeling epidemics, compartmental models are vital for studying infectious diseases by providing a way to analyze the dynamics of the disease spread over time. The most basic of such models is known as the SIR model, which groups the population into three compartments (susceptible, infectious, recovered) and has a simple flow where an individual moves from susceptible to infectious to recovered ([3]). An additional compartment called the exposed group, can be included to obtain the SEIR model ([18]). This model is better suited for diseases with a latent period, the time when an individual has contracted the disease but is unable to infect others.

Although the SEIR model is a more accurate portrayal of an infectious disease than the SIR model, as most infectious diseases have a latent period, one limitation of this model is that it assumes that when someone recovers from the disease, they are immune to it forever. This is unrealistic because one can lose their immunity over time. The SEIRS model is used to rectify this issue as this model assumes that individuals in the recovered group are able to return to the susceptible group ([4]).

Here we take this theme even further and introduce our SE(R)IRS model (Section 2), which generalizes the classical SEIRS model in two important ways. First, as the acronym suggests, individuals in the *E* compartment can pass directly to the *R* compartment without ever entering the *I* compartment. This choice was motivated by the recent COVID-19 pandemic, in which many people contracted the virus but recovered without ever demonstrating symptoms ([6, 14]). While it makes no difference mathematically when analyzing equilibria and their stability, here it is important to point out that because we take COVID-19 as our motivating example, in this paper we think of the *E* compartment as representing infected but asymptomatic individuals, while the *I* compartment represents individuals who are both infected and symptomatic. We have chosen to retain the traditional labeling for simplicity.

This naturally leads to our second generalization of the classical SEIRS model: we allow individuals from the *E* compartment to infect those who are susceptible, although at a lesser rate than those from the *I* compartment. This tracks with COVID-19, as asymptomatic individuals have been shown to transmit the disease, but less so than symptomatic individuals ([11, 19, 20]).

In Section 3 we generalize further still, adding a *V* compartment representing vaccinated individuals. The resulting model, which we call SVE(R)IRS, is significantly more complicated. We are still able to derive a few minor properties, but this section is an open invitation to future research. A novel feature is that one can treat the vaccination rate as a control (as it depends on people, not the disease) and subsequently consider some interesting optimal control problems.

Creating a model is one thing, while analyzing it is quite another. Here we have chosen to focus our analysis on the equilibria of the systems and their stability. This choice was also motivated by COVID-19 and what will be the “end” of the pandemic. As the concept of herd immunity has received much attention in both the media and academia ([1, 2, 5, 9, 21]), so far the notion of endemic equilibrium seems both important and relatively inconspicuous in the public discourse. There is a mathematical foundation for the idea of herd immunity ([13]), but as we demonstrate in Section 4.1, this does not mean the disease is eradicated. It is compatible with what we consider the more relevant idea of an endemic equilibrium: that the disease will always exist (hopefully in small enough numbers to no longer characterize a pandemic). Moreover, the stability of such an endemic equilibrium would reflect the possibility that new outbreaks or variants could cause spikes in infections, but that over time these numbers would drift back towards some state of “new normal.” The main idea is to design maintenance strategies to control the dynamic of the spread of the disease (through a yearly vaccine or seasonal non-pharmaceutical measures) to stay in a neighborhood of a sustainable endemic equilibrium.

While this discussion makes clear that our models and objects of study are motivated by COVID-19, we hope that our contributions can be applied to other infectious diseases with similar characteristics, including those yet to be discovered.

As in any mathematical modeling, there is naturally a trade-off between a model’s complexity and its accuracy. In many ways the classical SIR model is useful mainly due to its simplicity, making both mathematical analysis and simulations painless. But its accuracy may be consequently limited. On the other hand, much more complicated compartmental models, such as that in [8], may represent the dynamics of the disease very well at the expense of being computationally difficult. We hope that, like the popular SEIRS model, the models introduced here strike a reasonable balance by being simple enough for elementary dynamical systems theory and computations, while proving more flexible and accurate than the SEIRS model. In particular, our main result, Theorem 2.2, characterizes the stability of both the endemic and disease-free equilibria in terms of the basic reproductive number ℜ_0_ using only basic theory. But ignoring the effects of vaccination greatly misrepresents the course of pandemics like COVID-19. Yet our SVE(R)IRS model, while more accurate, was just complicated enough that similar analyses failed and we could prove no such theorem. For these reasons we believe these models lie somewhere near the right balance of complexity and simplicity. To our knowledge, neither has appeared in the literature before.

## 2. SE(R)IRS model

The usual SEIRS model ([4]) can be visualized as

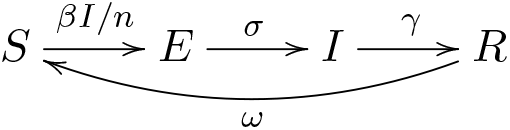

where

- *β* is the transmission rate, the average rate at which an infected individual can infect a susceptible
- *n* is the population size
- 1*/σ* is the latency period
- 1*/γ* is the symptomatic period
- 1*/ω* is the period of immunity.

All parameters are necessarily non-negative. Note that, for simplicity, here and throughout this paper we choose to present our models without vital dynamics (also known as demography), often represented by the natural birth and death rates Λ and *µ*. Also note that when *ω* = 0, this reduces to the usual SEIR model, and one can further recover the simple SIR model by letting *σ → ∞*.

As described in Section 1, we now think of individuals in the *E* compartment as infected but asymptomatic, while individuals in the *I* compartment are infected and symptomatic. Therefore in our new SE(R)IRS model certain asymptomatic individuals can recover without ever becoming symptomatic; the duration of the course of their infection is denoted by 1*/δ*. Moreover, asymptomatic individuals can indeed infect susceptible individuals, however they do so at a reduced rate when compared to symptomatic individuals; this reduction is accounted for by the parameter *α*. We may assume *α* ∈ [0, 1] and *δ* ≥ 0. Note that when *α* = *δ* = 0 we recover the SEIRS model.

The SE(R)IRS model can be visualized as

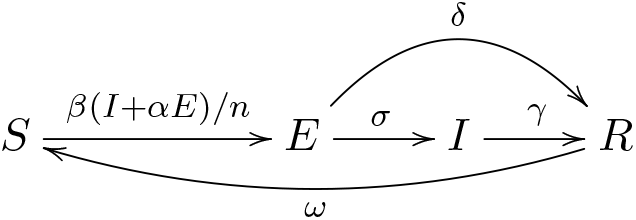

and the corresponding dynamical system is given by

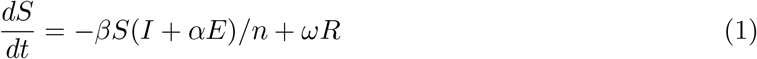

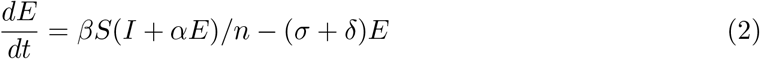

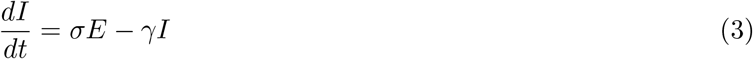

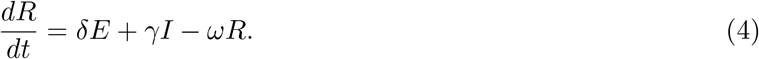

In any of these various compartmental models, the equilibrium points come in two types. A disease-free equilibrium has no individuals in the *E, I*, or *R* compartments; this represents a steady state where there is no disease at all. Any other equilibrium point is considered an endemic equilibrium; this represents a constant state where there is always some proportion of the population infected by the disease. The first step towards analyzing either type of equilibrium is to calculate the basic reproductive number, which represents the average number of cases directly resulting from a single infection in a population of only susceptible individuals.

### Proposition 2.1.

*The SE(R)IRS basic reproductive number is*

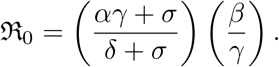

*Proof*. We follow [12] by computing ℜ_0_ as the spectral radius of the next generation matrix. First, it is easy to see that the system has a disease-free equilibrium at (*S, E, I, R*) = (*n*, 0, 0, 0). We compute

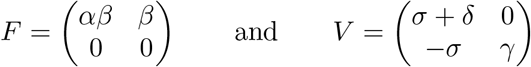

so our next generation matrix is

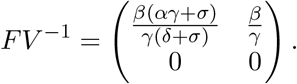

The basic reproductive number ℜ_0_ is the spectral radius of this operator, which is the largest eigenvalue 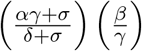.

If we use the fact that *n* = *S* + *E* + *I* + *R* is constant we can reduce this to a 3 × 3 system in *S, E, I*. The reduced equations are

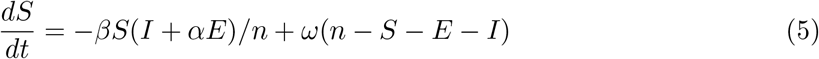

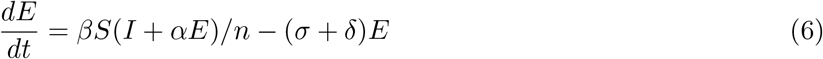

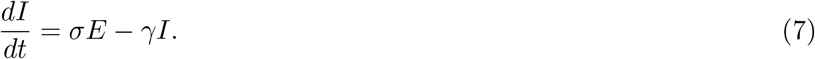

In the sequel we will study this simpler version of the system, recovering the value of *R* when convenient.

Our main result, proved in the next two sections, is the following.

### Theorem 2.2.

*If* ℜ_0_ *<* 1 *then the disease-free equilibrium is locally asymptotically stable and the endemic equilibrium is irrelevant. If* ℜ_0_ *>* 1 *then the endemic equilibrium is locally asymptotically stable and the disease-free equilibrium is unstable*.

*Proof*. The theorem follows from Lemmas 2.3, and 2.4, and 2.7.

Here “irrelevant” means epidemiologically nonsensical, as certain compartments would contain negative numbers of people; it still exists mathematically. This theorem is sometimes known as the “endemic threshold property”, where ℜ_0_ is considered a critical threshold. In [13], Hethcote describes this property as “the usual behavior for an endemic model, in the sense that the disease dies out below the threshold, and the disease goes to a unique endemic equilibrium above the threshold.” The SEIR version is derived nicely in Section 7.2 of [18]. The SEIRS version can be found in [17].

### 2.1. Analysis of endemic equilibria

Our system has a unique endemic equilibrium at

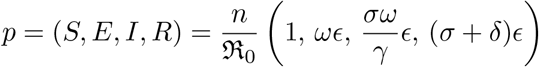

where

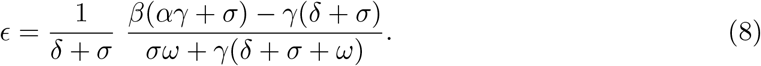

Note that this endemic equilibrium is only realistic if all coordinates are positive, which requires *ϵ* positive, which requires

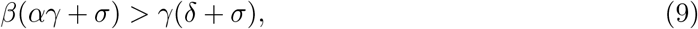

which is equivalent to

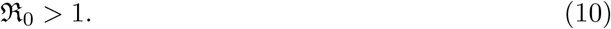

In other words, we have the following.

#### Lemma 2.3.

*If* ℜ_0_ *<* 1 *then the endemic equilibrium contains negative coordinates, and is thus epidemiologically irrelevant. If* ℜ_0_ *>* 1 *then all coordinates are positive*.

The linearization of the reduced system at *p* is

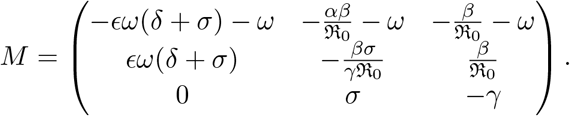

As expected, this matrix does not depend on *n*. Unfortunately, the eigenvalues of *M* are not analytically computable for general parameters. Note that *M* is nonsingular for generic parameter values. But the determinant does indeed vanish if and only if ℜ_0_ = 1.

#### Lemma 2.4.

*If* ℜ_0_ *>* 1 *then the endemic equilibrium is locally asymptotically stable*.

*Proof*. We apply the criteria (12.21-12.23) from [10] to *M*. Note that the trace is obviously negative, so (12.22) is immediately satisfied. Now compute

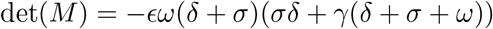

which is clearly negative, so (12.21) is satisfied. Finally, we compute the bialternate sum of *M* with itself,

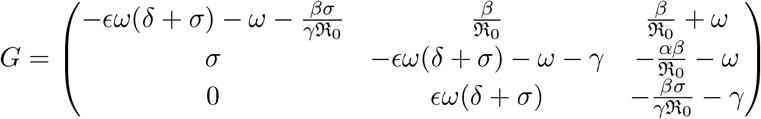

whose determinant

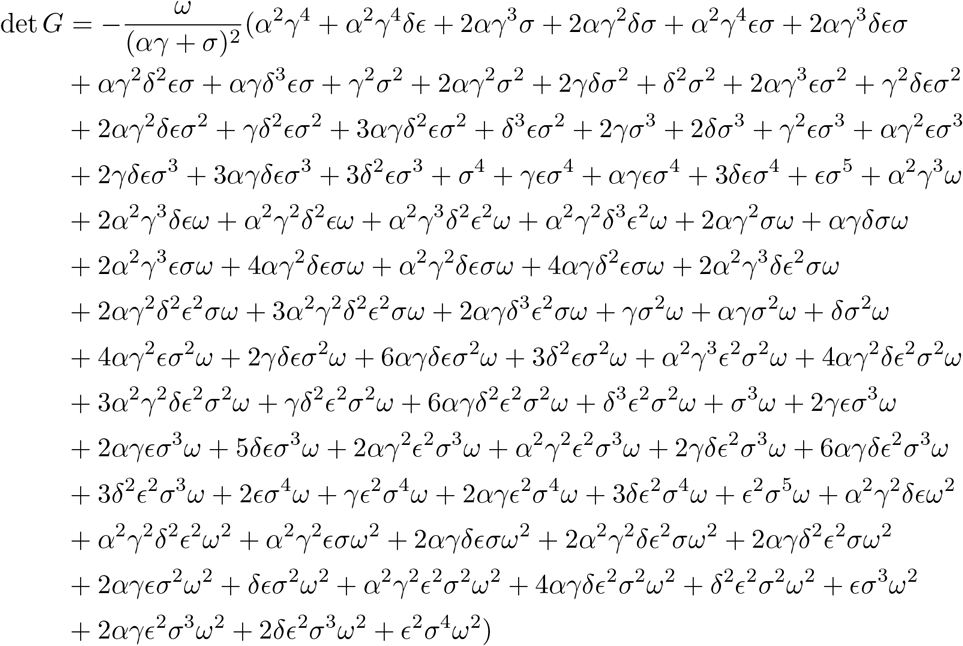

is also negative, satisfying (12.23).

#### Example 2.5.

In all of our examples the time units are taken to be days, and we choose the parameter values

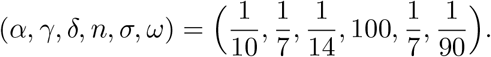

These values are somewhat realistic for COVID-19. While there are no known precise values, these are at least roughly in agreement with some of the literature. Specifically, we assume that an asymptomatic individual is 10% as infectious as a symptomatic one, that individuals who become symptomatic have seven day periods of latency and of symptoms, that individuals who never develop symptoms are infected for 14 days, and that the period of immunity is 90 days. We choose the population size of 100 simply so that compartment values can be interpreted as percentages of a generic population.

When *β* = 0.4 we have

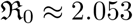

and

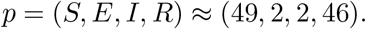

The eigenvalues of *M* are

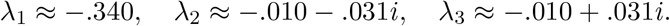

These three eigenvalues all have negative real part, so the equilibrium is stable. It appears to be a spiral sink, signifying epidemic waves ([4]), as shown in Figure 1. In Figure 2 we see that for small numbers of initial infections, the number of individuals in the *E* and *I* compartments initially rise, before settling down toward the smaller values of the endemic equilibrium. This may hold a lesson for officials and policy-makers: an initial spike in cases (perhaps caused by a new variant) does not always portend an exponential outbreak necessitating intervention. With patience the case numbers may naturally drop back to the endemic equilibrium.

This example represents a potentially new normal post the COVID-19 pandemic, with relatively small but nonzero proportions of the population infected at any given time.

**Figure 1.**
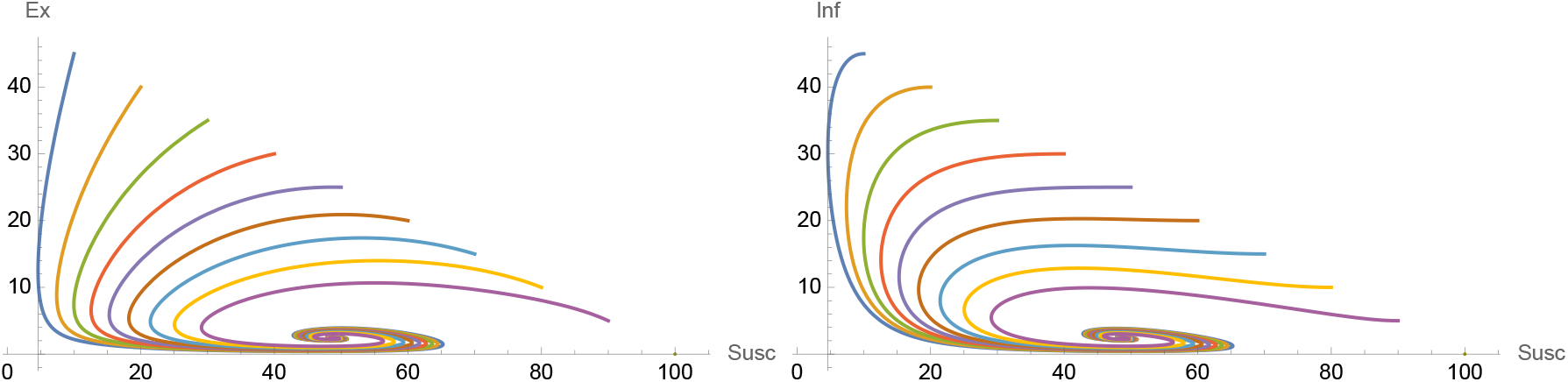
The endemic equilibrium of Example 2.5. The left plot displays *E* versus *S* compartments, while the right plot displays *I* versus *S* compartments. Each colored curve represents a different initial condition of the form (*S,E,I,R*) = (*S*_0_, (*n* – *S*_0_)/2, (*n* – *S*_0_)/2, 0) for *S*_0_ = 10, 20,…,90.

**Figure 2.**
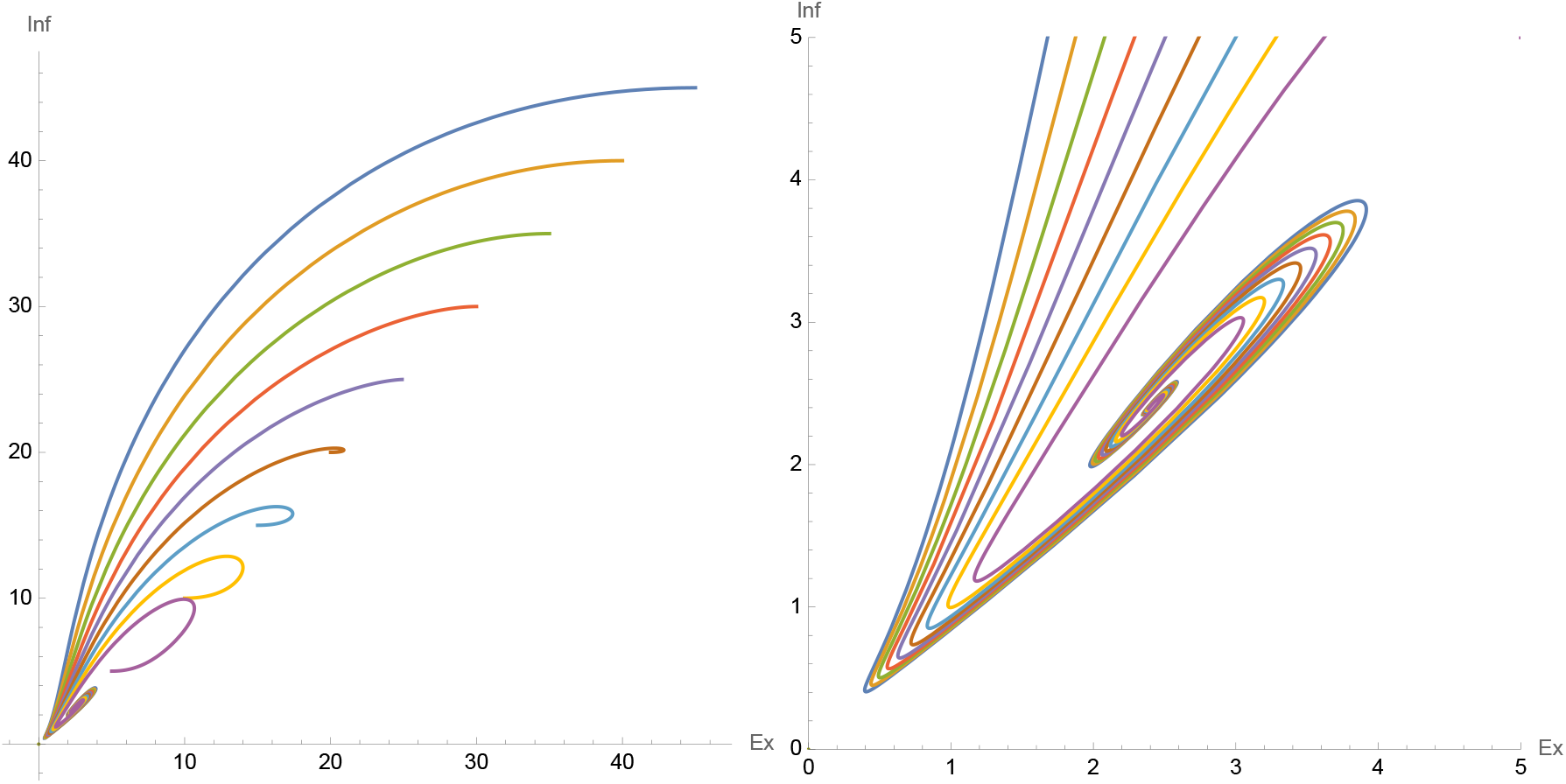
The endemic equilibrium of Example 2.5 with colors as in Figure 1. The left plot displays the *I* versus *E* compartments. The right plot shows the same, zooming in on the equilibrium point.

#### Example 2.6.

Taking *β* = 0.2 and the other parameters as in the previous example yields

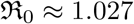

and the endemic equilibrium

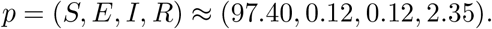

In this relatively realistic scenario, on any given day we find 97.40% of our population is susceptible, 0.12% is asymptomatic, 0.12% is symptomatic, and 2.35% is recovering yet immune.

### 2.2. Analysis of disease-free equilibria

One easily checks that the system has a disease-free equilibrium at (*S, E, I, R*) = (*n*, 0, 0, 0). The linearization of the reduced system there is

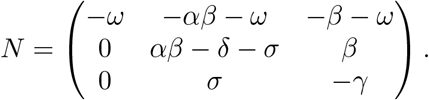

This is singular if and only if ℜ_0_ = 1, just like *M*.

#### Lemma 2.7.

*The disease-free equilibrium is locally asymptotically stable if and only if* ℜ_0_ *<* 1.

*Proof*. The eigenvalues of *N* are

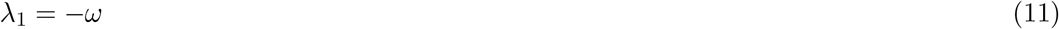

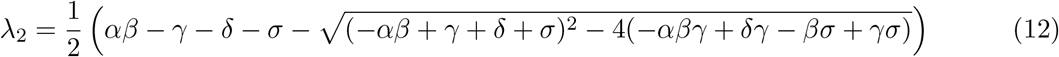

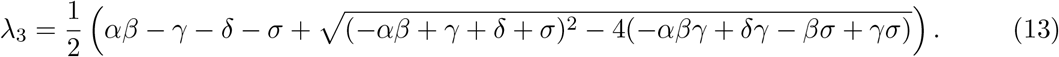

It is not obvious, but algebra shows that all three eigenvalues are real since our parameters are positive: the discriminant simplifies to 4*βσ* + (*αβ* + *γ − δ − σ*)^2^. Now *λ*_1_ is clearly always negative. Next, we have

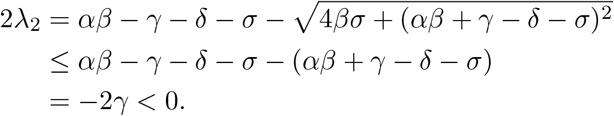

Thus *λ*_2_ is also always negative, and in fact we have

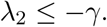

Finally, Mathematica shows that

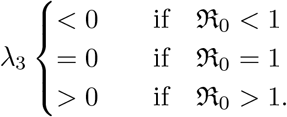

Thus (*n*, 0, 0) is stable if and only if ℜ_0_ *<* 1.

#### Example 2.8.

When

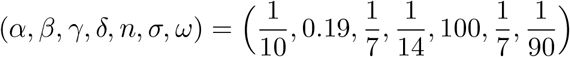

we have the disease-free equilibrium at (*S, E, I, R*) = (100, 0, 0, 0) and

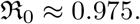

The eigenvalues of *N* are

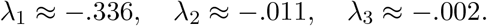

The three eigenvalues are negative real, so the equilibrium is stable. See Figure 3.

**Figure 3.**
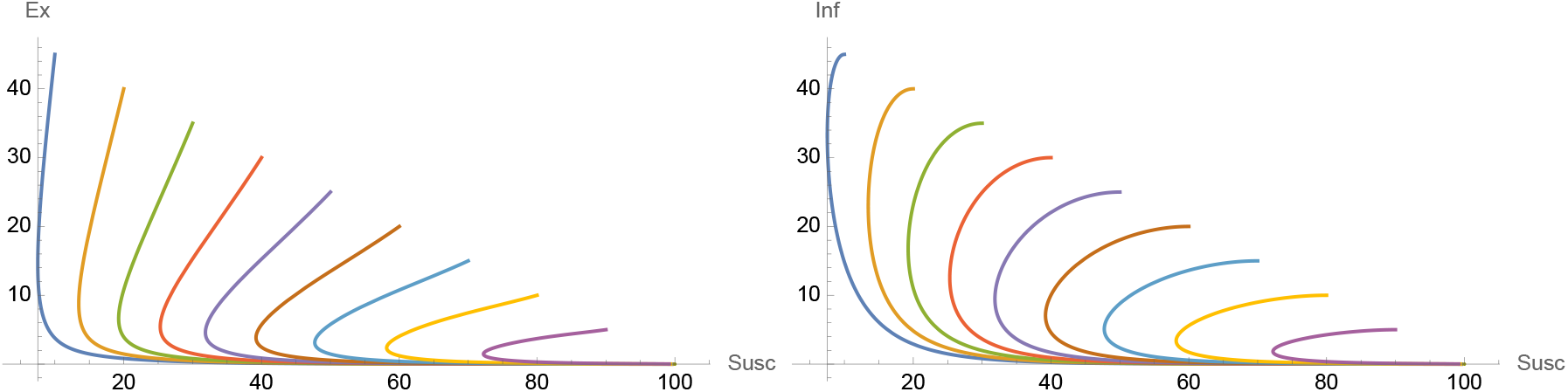
The disease-free equilibrium of Example 2.8. Plots as in Figure 1.

## 3. SVE(R)IRS model

Adding a vaccinated compartment to the model in Section 2 yields the following model, *which we denote SVE(R)IRS:*

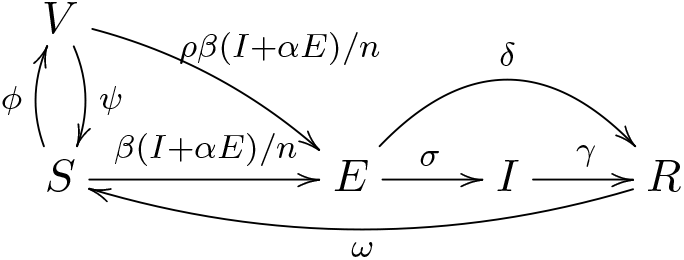

Here 1 – *ρ* represents the efficacy of the vaccine, 1*/ψ* is the duration of efficacy of the vaccine, and 1*/ϕ* is the rate at which people are vaccinated. The first two are intrinsic to the vaccine itself, while *ϕ* can be thought of as a control (see Section 4.2). We may assume *ρ* ∈ [0, 1] and *ϕ, ψ >* 0.

The associated dynamics are given by:

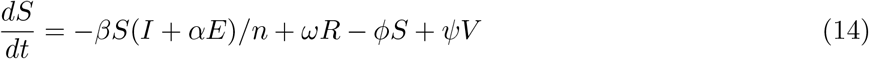

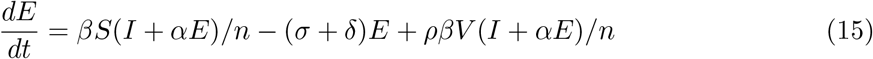

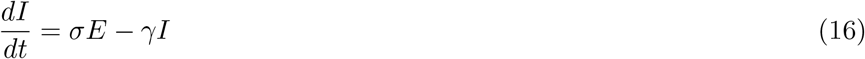

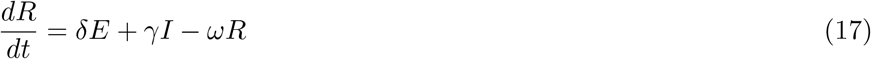

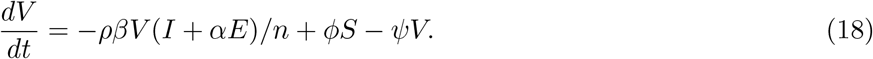

The dynamics of this model are significantly more complicated than those of the SE(R)IRS model in the previous section. We do manage to prove the analogue of Lemma 2.7 holds. We were unable to prove the analogues of Lemmas 2.3 and 2.4, although experimental evidence suggests that both hold.

### Proposition 3.1.

*The SVE(R)IRS basic reproductive number is*

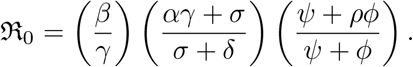

*Proof*. Again we follow [12] by computing ℜ_0_ as the spectral radius of the next generation matrix. We use tildes to not confuse with the compartment *V*. First, a short computation shows that the system has a disease-free equilibrium at 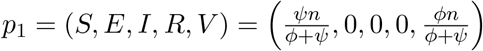. We compute

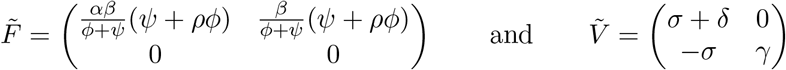

so our next generation matrix is

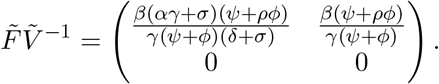

The basic reproductive number is the spectral radius of this operator, which is the largest eigenvalue: 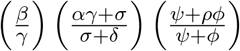.

If we use the fact that *n* = *S* + *E* + *I* + *R* + *V* is constant we can reduce this to a 4 × 4 system in *S, E, I, V*. The reduced equations are

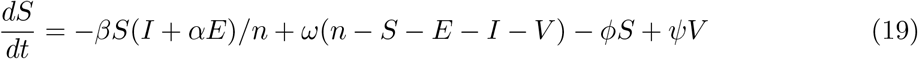

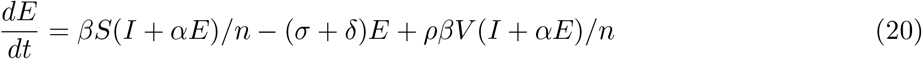

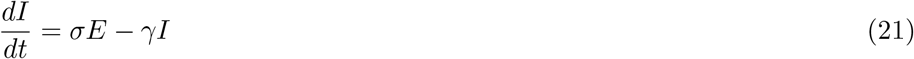

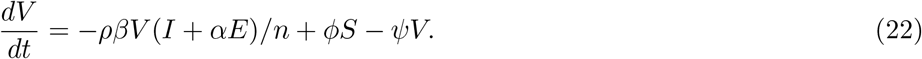

In the sequel we will study this simpler version of the system, recovering the value of *R* when convenient.

### 3.1. Analysis of endemic equilibria

Mathematica gives two endemic equilibria, *p*_2_ and *p*_3_, which are square root conjugate to each other. One of them is given by

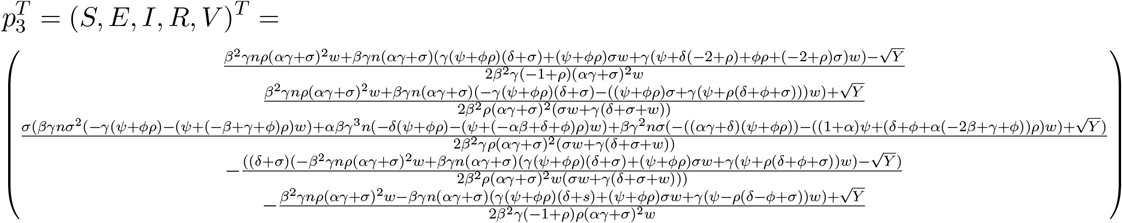

where

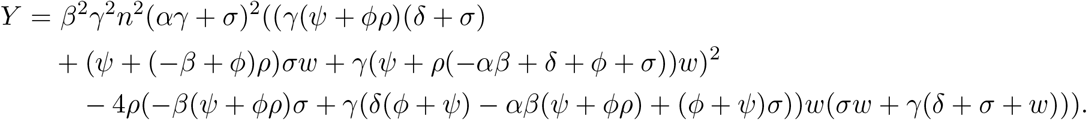

These are clearly very complicated, and it is not clear whether the components are even positive for positive parameters. Examples suggest that for realistic COVID-19 parameters, *p*_2_ has some negative components and can be ignored, while *p*_3_ has all positive components and should be treated as a realistic endemic equilibrium.

The linearization of our system at either point is unwieldy; Mathematica cannot even determine when the determinant vanishes, let alone compute eigenvalues. It can, however, produce the characteristic polynomial, so the methods of [10] applied in the proof of Lemma 2.4 could potentially work. However, the characteristic polynomial is a complex expression and it is hard to tell whether the coefficients are positive; something similar is expected for *G*, the bialternate product of this matrix with itself.

Therefore the existence, uniqueness, and stability of *p*_3_ remain open. The methods used to prove Lemmas 2.3 and 2.4 could potentially work with more insight or computational power, but other techniques might be necessary. For now we limit ourselves to examples, which provide hope that the analogues of these Lemmas may indeed hold for the SVE(R)IRS model.

#### Example 3.2.

When

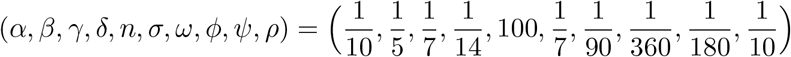

we have ℜ_0_ ≈ 0.719 and we find that neither *p*_2_ nor *p*_3_ contains all positive coordinates. So there is no relevant endemic equilibrium for these parameters. Section 3.2 shows that there is in fact a stable disease-free equilibrium.

If we free *ϕ* and keep all other parameters the same, we find that *p*_3_ has all positive coordinates if and only if *ϕ <* 0.000165, which unsurprisingly corresponds precisely to those *ϕ* values for which ℜ_0_ *>* 1.

#### Example 3.3.

Here we keep all parameters the same as in Example 3.2 except *β*. When

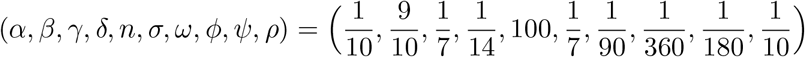

we have

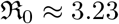

and an endemic equilibrium at

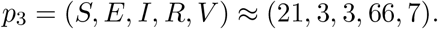

The point *p*_2_ contains negative components and is thus irrelevant.

The eigenvalues of the reduced linearization at *p*_3_ are

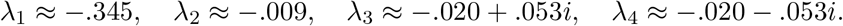

The four eigenvalues all have negative real parts, so the equilibrium is stable. Note, however, that they are not all real, in contrast to the disease-free equilibrium case (see Proposition 3.4 below). We again see the appearance of a spiral sink due to epidemic waves ([4]). See Figure 4.

**Figure 4.**
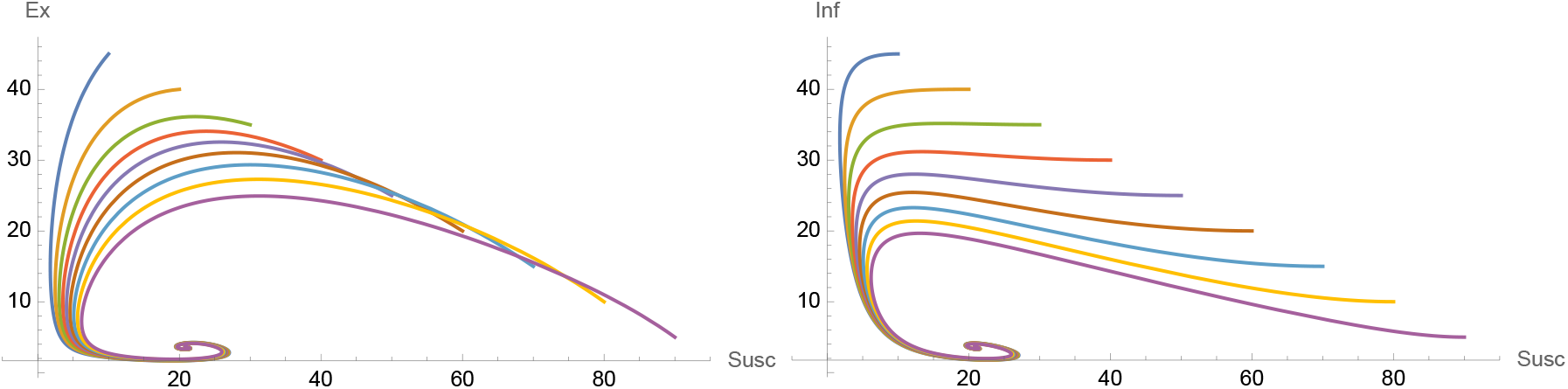
The endemic equilibrium of Example 3.3. Plots as in Figure 1.

### 3.2. Analysis of disease-free equilibrium

Setting *E* = *I* = 0 and solving the resulting system yields the unique disease-free equilibrium

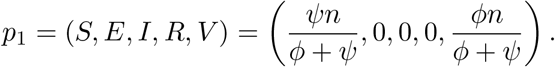

Linearizing at this point yields the matrix

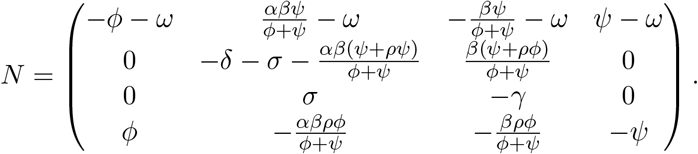

As in the SE(R)IRS model, this matrix is singular if and only if ℜ_0_ = 1.

#### Proposition 3.4.

*The disease-free equilibrium is locally asymptotically stable if and only if* ℜ_0_ *<* 1.

*Proof*. The eigenvalues of *N* are

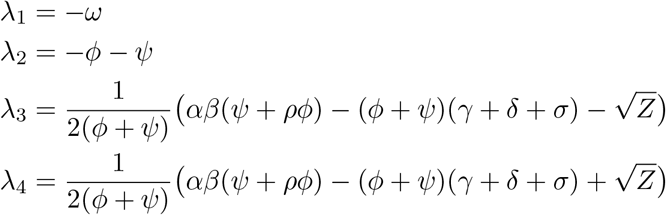

where large amounts of tedious algebra show that

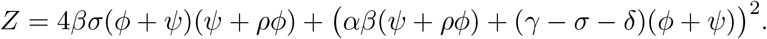

This form of *Z* makes it apparent that *Z* is always positive and thus all four eigenvalues are always real.

Now it is also clear that *λ*_1_ and *λ*_2_ are always negative. Next, we have

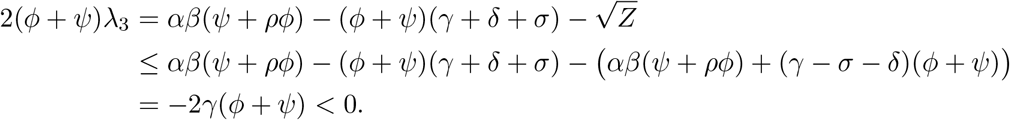

Thus *λ*_3_ is always negative as well, and in fact we have

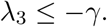

Finally, Mathematica shows that det *N >* 0 if and only if ℜ_0_ *<* 1. Since det *N* = *λ*_1_*λ*_2_*λ*_3_*λ*_4_ and *λ*_1_, *λ*_2_, *λ*_3_ *<* 0, this shows that *λ*_4_ *<* 0 if and only if ℜ_0_ *<* 1. This proves the Proposition.

## 4. Discussion

In this last section we address the question of herd immunity versus endemic equilibrium, and we open a discussion of optimal strategies for vaccination. We end conclude by stating some open problems.

### 4.1. Herd immunity

In addition to our perceived need to consider compartmental models more closely adapted to COVID-19 than the classical models, this work was motivated in part by our observation that the national dialogue concerning the post-pandemic future focused largely on the concept of herd immunity rather than that of endemic equilibria. According to [13], one has herd immunity when the immune fraction of the population exceeds 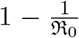. In this case, the disease “does not invade the population”. However, for classical models as well as SE(R)IRS, we know that as long as ℜ_0_ *>* 1 we still do not reach a disease-free equilibrium which would correspond to the complete eradication of the disease.

#### Example 4.1.

To illustrate the shortcomings of the herd immunity concept, reconsider Example 3.3 which includes annual vaccinations. Then we have 1 – 1/ℜ_0_ ≈ 0.69. According to [13], herd immunity thus occurs if more than 69% of the populations is immune to the disease. However, at the endemic equilibrium for these parameters, we have *R* ≈ 66 and *V* ≈ 7. Thus 73% of the population is immune, which is above the required threshold. But over 6% of the population is either in the *E* or *I* compartment (≈ 3.4% each). This shows that although we have technically achieved herd immunity, large numbers of people still suffer from the disease.

#### Example 4.2.

Ideally, sufficient vaccination could eradicate a disease completely. By Proposition 3.4, this could happen if we took *ϕ* large enough to force ℜ_0_ *<* 1, as the dynamics would trend toward the disease-free equilibrium. However, consider the following parameters, which are realistic for COVID-19:

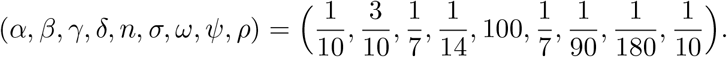

Here we have left *ϕ* free as a control. We compute that ℜ_0_ *<* 1 if and only if *ϕ >* 1*/*282. Thus, in order to set a trajectory towards disease eradication, the population would need to be revaccinated more often than once per year, which seems unlikely. Therefore for these parameters it seems more realistic that the best we could hope for is an endemic equilibrium with relatively small portion of the population infected at any given time. With annual vaccinations we would have approximately 0.73% of the population in the *E* or *I* compartments.

### 4.2. Strategies for Vaccination

The vaccine can be thought of as a control over the system to steer the variables to desired values. We can express our model as an affine control system with drift as follows. Let *u* = *ϕ* be our control and *q* = (*S, E, I, R, V*)^*T*^ be our state. Then the dynamics are given by

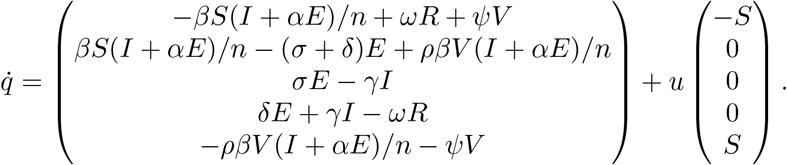

The first vector field here is the drift, and the second is the control vector field.

If we fix a time period *T* we have several meaningful choices of cost function *J*(*u*), obtaining an optimal control problem. For example, we could choose *J* to be the final number of symptomatic individuals *I*(*T*), or the total number of symptomatic individuals over the period 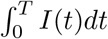, or the number of symptomatic individuals at the endemic equilibrium (if it exists and is unique). Note that the number of symptomatic people can be considered a proxy for the number of hospitalizations or deaths. Then seeking the optimal vaccination strategy for minimizing illness or hospitalizations or deaths can be modeled as minimizing *J*(*u*) subject to the dynamics (19) – (22). Forthcoming work will utilize tools from geometric optimal control to determine the existence and role of singular arcs in optimal strategies.

However, initial numerical simulations have given somewhat uninteresting, if realistic, optimal strategies. We used the optimal control software Bocop ([22]) to simulate the optimal control *u*(*t*) = *ϕ*(*t*) for a small sample of initial conditions, bounds, and cost functions. These experiments suggest that the optimal vaccination strategy is to constantly vaccinate at the maximum rate for the costs 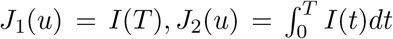, and 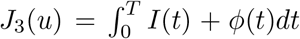. Using the 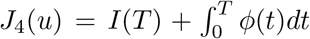 yielded the optimal strategy of vaccinating nobody until near the end of the period *T*, then vaccinating at the maximum rate for a short burst. While these strategies make sense, they are not particularly mathematically interesting. However, at this point we only have a small sample of numerical results; a rigorous control theoretic investigation is needed. Unfortunately our software was not able to handle the cost function given by the *I*-component of the endemic equilibrium *p*_3_ (see Section 3.1), but this formulation of the problem could potentially yield results which are interesting both mathematically and epidemiologically.

### 4.3. Open questions

There remain a large number of open questions suggested by this work as well as directions for generalization. The most obvious question is whether the endemic threshold property in Theorem 2.2 holds for the SVE(R)IRS model of Section 3. It may be that one, both, or neither of the analogues of Lemmas 2.3 and 2.4 hold for SVE(R)IRS. In fact, in the SVE(R)IRS model both the existence and the uniqueness of the endemic equilibrium remains to be established in the ℜ_0_ *>* 1 case. All the examples we have explored suggest that the answers to these questions are affirmative, but with so many free parameters an exhaustive search for counterexamples is challenging.

A different set of open questions concerns generalization of both our models and our results. In particular, a natural addition to either model would be the vital dynamics of birth and death rates, as is common in the SEIRS literature. We suspect that this would not qualitatively affect our results (especially in the fixed population case), but made no attempts at studying these problems. Other generalizations could include nonlinear transmission or seasonal forcing, as well as the appearance of new variants which would translate in a time varying coefficients such as *β*. An alternative research direction would be the study of global rather than local stability of equilibria. Global stability for classical compartmental models has an extensive literature; see the comprehensive review article [13]. In particular, global stability was treated in [15] and [17] for SEIR, and in [7] and [16] for SEIRS. The methods of these and related papers might also prove effective in our models.

## Data Availability

n/a

